# Determinants of Placenta Previa among Pregnant Women Delivered in Public Hospitals South Ethiopia: Unmatched Case-Control Study

**DOI:** 10.1101/2025.02.02.25321556

**Authors:** Tadiwos Utalo, Eskinder Wolka, Tagese Yakob Barata, Awoke Abraham, Getu Kassaye, Jinenu Getu

## Abstract

**Background:** Placenta previa is one of the serous obstetric complications in which the placental tissue abnormally implants the lower uterine segment partially or totally. Although the exact etiology of placenta previa is unknown, certain factors can contribute to the condition. This study was aimed to identify the determinants of placenta previa among pregnant women delivered in public hospitals in the wolaita zone, southern regions of Ethiopia.

**Methods:** A facility-based unmatched case-control study was conducted on 144 cases and 287 controls from July 8, 2020 to June 30, 2023. The sample size was determined using Epi Info™7.2.5.0. Study subjects were selected using consecutive sampling method. Data were collected using structured checklists, coded, and then input into Epi-data manager version 4.6.0.2 for analysis using SPSS version 25. Using both bivariate and multivariable logistic regressions, the determinants of placenta previa in pregnant women were identified. Adjusted odds ratio with 95% confidence interval and p-value <0.05 were used to declare the significant association.

**Results:** The clinical data of 274 pregnant women with placenta previa group and 388 pregnant women with normal placental position (general group) who delivered in our hospital from July 8, 2020 to June 30, 2023 were retrospectively analyzed. Out of 19,677 hospital deliveries, 144 pregnant women with placenta previa (cases) and 287 without placenta previa (controls) were included in this study. The mean age of the mothers was 27.35±5.04 years for both cases and controls. The odds of developing placenta previa is higher for the maternal age of ≥35 years[AOR=4.45 (95% C.I =1.2, 4.99)], short inter-pregnancy interval of less than 24 months [AOR=1.89 (95% C.I=(1.08, 3.53)], previous abortion [AOR=5.49 (95% C.I =2.93, 10.3)], previous cesarean sections [AOR=3.73 (95% C.I =1.68, 8.3)], and women having uterine leiomyoma [AOR=6.33 (95% C.I=2.48, 16.17)].

**Conclusion:** Increased maternal age (≥35 years), short inter-pregnancy interval of less than 24 months, history of abortions, prior cesarean sections, and having leiomyoma were identified determinants of placenta previa.

## Introduction

Placenta previa (PP) refers to abnormal implantation of placental tissue to the lower uterine segment either overlies or close to internal cervical os. Currently, pacenta preveia categorized as placenta previa (when placental edge is overlie the internal os totally or partially), and low lying placenta (when the placental edge is either reaches the margin or within 2 cm of internal cervical os) [1-3]. PP is one of the serous causes of excessive obstetric hemorrhage during pregnancy and childbirth. It may result in life-threatening complications, such as hemorrhage before, during or after childbirth, cesarean hysterectomy, or surgical complications [3]. The exact cause of placenta previa is unknown, but previous research shows there are determinants for the development of placenta previa. Some of the determining factors for PP were advanced maternal age (above 34 years), multi-parity, a previous Caesarean section, a history of abortion or uterine curettage, cigarette smoking, previous placenta previa, and multiple pregnancies and male sex [4-9].

Globally, the prevalence of placenta previa is 3.5 per 1000 pregnancies, and it is still increasing due to the increased trend of operative delivery ([10]. The development of PP varies from region to region due to a difference in risk factors. The prevalence was highest among the Asian population (12.2 per 1000 pregnancies and lower among Sub-Saharan Africa (2.7 per 1000 pregnancies) [11]. In Ethiopia, the prevalence of placenta previa was 0.7% [4]. Globally, obstetric hemorrhage is one of the leading causes of maternal death, including Sub-Sahara Africa (SSA) which accounts for 24.5% in SSA [12]. According to the Ethiopian national maternal death surveillance and response (MDSR) report of 2010EFY, obstetric hemorrhage was the leading cause of maternal mortality; accounts for 41.3 % of all causes of maternal death, and 76% of these deaths were due to PPH [13]. Placenta previa accounts about more than 10% of neonatal mortality and 0.7% of maternal mortality [14, 15]. Ante-partum hemorrhage (APH) complicates 2-5% of all pregnancies which has various causes, up to one-third are due to placenta previa [16]. Globally, Postpartum hemorrhage (PPH) is one of the leading causes of maternal death and about 14 million women develop PPH resulting in 20% (about 70,000) of maternal death annually despite being preventable and treatable [17].

The feared complications of placenta previa are hemorrhage and shock in the mother and preterm delivery (before 37 weeks), Intra-uterine growth restriction (IUGR), stillbirth, and neonatal death in the neonate. It contributes about 27.4% for the occurrence of postpartum hemorrhage [18]. Placenta previa is also associated with increased Caesarean section and placenta accreta spectrum (PAS) pathologies such as placenta accreta, placenta increta, and placenta percreta), which can lead to Caesarean hysterectomy, septicemia, intensive care unit (ICU) admission, thrombophlebitis, and even maternal death [19, 20].

For pregnant women with placenta previa, perinatal mortality rates are three to four times higher than in those without placenta previa pregnancies [21]. Despite of the bad effects of the PP during pregnancy, postpartum, and also in subsequent pregnancies, very little was known about the determinants of placenta previa among pregnant women in Ethiopia in general, and there was no research finding in this study area particularly. To avert the placenta previa related pregnancy and child birth complications for both the mother and newborn, identifying the risk factors of PP is far most important in contributing safe journey of pregnancy and improving feto-maternal outcomes. Hence, this study was aimed to identify the determinants of placenta previa among pregnant women delivered after 28 weeks’ of gestational age in public hospitals in the wolaita zone, southern regions of Ethiopia.

## Methods and Materials

### Study setting, design, and Period

A facility-based unmatched case-control study was conducted from August 1, 2023, to August 30, 2023 in three selected public hospitals in the wolaita zone southern regions of Ethiopia among all pregnant women with the diagnosis of placenta previa (cases) and those without PP (controls) who admitted and managed from July 8, 2020 to June 30, 2023. Wolaita zone located in newly emerged south regional state of Ethiopia. Wolaita sodo, capital city of Wolaita zone is situated 330 Km South of Addis Ababa, Ethiopia’s capital city. Zone has 25 districts with an estimated total population of more than 4 million. It has 9 public hospitals (one comprehensive specialized hospital and 8 primary hospitals) during this study period. These hospitals provide health services for more than 7.5 million of its total catchment populations [22].

### Populations

The source population of this study was all pregnant women who delivered in public hospitals from July 8, 2020 to June 30, 2023 whereas the study population included in this study was all pregnant women delivered after 28 weeks’ of gestation in selected public health facilities with diagnosis of placenta previa by ultrasound or during Caesarean sections from July 8, 2020 to June 30, 2023, were taken as cases and those pregnant women delivered without the diagnosis of PP were included as controls.

#### Inclusion and Exclusion Criterion

**For Cases:** all pregnancies diagnosed with placenta previa by sonography (all types) which confirmed by obstetrician’s or radiologist, or during Caesarean sections (intra-operative finding) after 28weeks of gestational age were included [23].

**For Controls**: all pregnancies with no diagnosis of placenta previa during delivery after 28weeks of gestational age were included. Whereas:

**For cases** out of 274 cases, 36 charts with abruptio placenta, 24 charts with heavy shows, 28 records later on diagnosed as abortions (<28 weeks), 20 charts with other diagnosis and 22 women with incomplete medical records about demographic characters, obstetric characteristics and clinical factors were excluded from the study and

**For controls**:-out of 388 women medical records, 42 charts with placenta abruption, 27 charts later on diagnosed as abortions (<28 weeks), 4 records with other diagnosis and 26 charts with incomplete and missed medical records were also excluded from the study.

#### Sample Size Determination

Double population proportion formula was used to estimate the sample size using Epi Info™7.2.5.0 version 3 considering the following statistical assumptions, Confidence interval =95%, Power of study=80%, Control to case ratio=2 to 1, percentage of controls exposed to risk factor (abortion in this case) = 18.5%, percentage of cases exposed to risk factor exposed (has abortion in this case) = 31.01% [4]. Sample size calculation was done for each determinant factor for PP and the one with the maximum sample size was selected for this study. Accordingly, a total of 431 pregnant women delivered after 28 weeks’ of gestational age were included in this study and from which 144 pregnant women diagnosed with PP and delivered after 28 weeks of gestational age by ultrasound or during Caesarean sections by seniors or resident were taken as cases, whereas, those 287 pregnant women without PP during any mode of delivery after 28 weeks of gestational age were included as controls.

#### Sampling Procedures

Initially, one comprehensive specialized hospital was purposely selected and two primary hospitals were selected randomly. Then, Health Management and Information System (HMIS) data was used to identify a total delivery. There was 13,138 total deliveries and 190 placenta previa group (cases) were identified during three year’s period in Wolaita Sodo university comprehensive specialized hospital, while a total of 3521 deliveries and 45 placenta previa group (case) were identified in Bele primary hospital for three year periods and a total of 3018 women gave birth and the total number of placenta previa case was 39 for three year periods in Tebela primary hospitals from July 8, 2020 to June 30, 2023 [24].

Then, emergency obstetric triage register and delivery register books were used to identify the admission and discharge records. Based on proportional allocation among total deliveries, 431 pregnant women were identified as eligible study populations, and from which 144 were cases and 287 were controls (**Figure 1**). The diagnosis of placenta previa (all types) was based on the ultrasound during admission and/ or during Caesarean sections by seniors. Consecutive sampling technique was conducted to select both the cases and controls. For each one case, two controls were selected consecutively (Case to control ratios=1:2) until calculated sample size was reached. A medical record about patients’ demographic (age, residence, occupation), obstetrics and clinical factors of the pregnant women was carefully identified.

**Fig 1:**
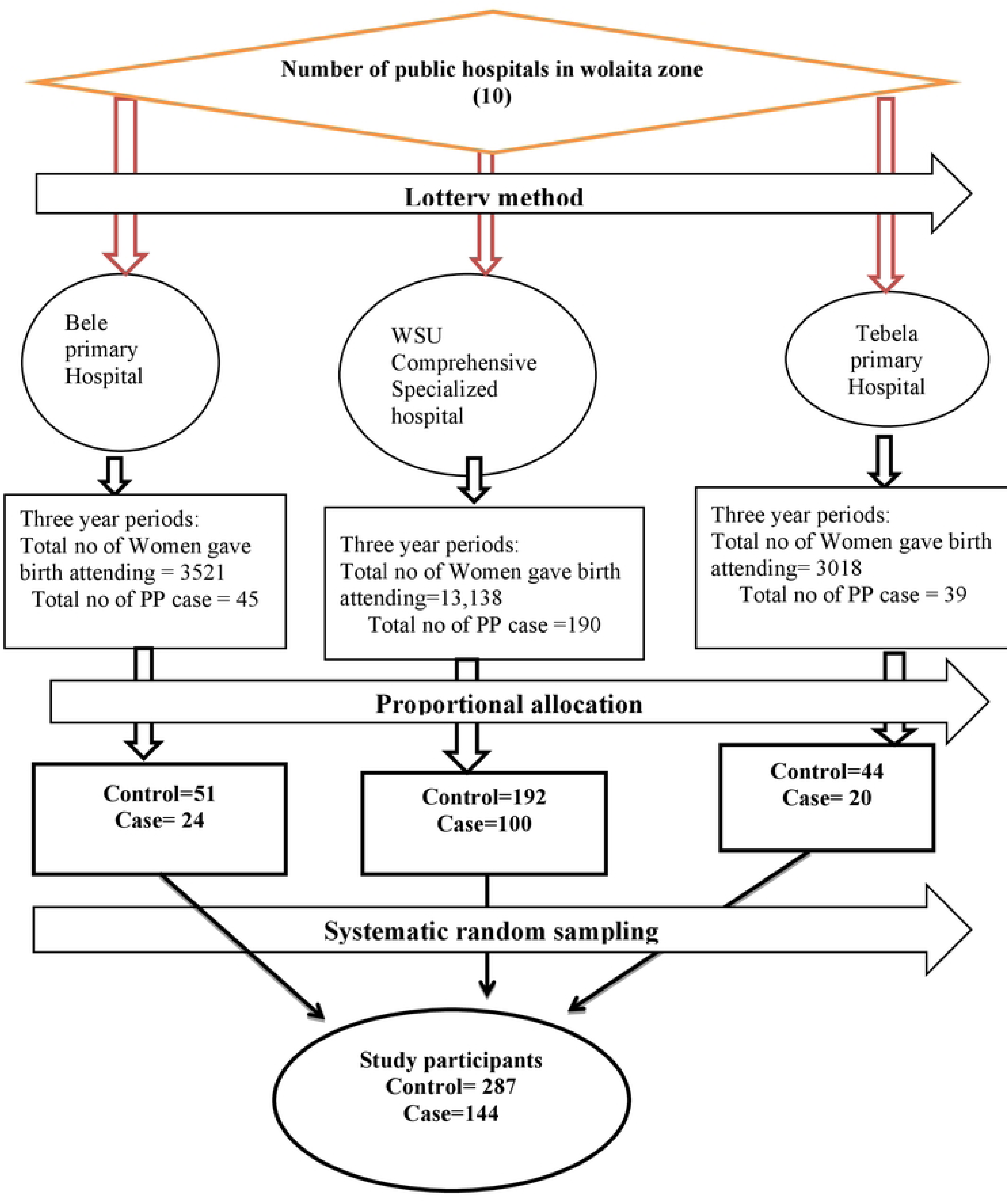
Diagrammatic presentation of the sampling procedure among pregnant women delivered public hospital across in the Wolaita zone Southern Ethiopia, 2023.

#### Study Variables

Independent Variables includes socio-demographic factors (age of the mother, marital status of the mother, residence, occupation of the mother), obstetric history (gravida, parity, months since last delivery, ANC, gestational age during ANC, obs U/S performed and type of U/S), obstetric characteristics(previous history of abortion, number of E and C, previous history of C/S, number of previous C/S, previous myomectomy, history of preterm delivery(before 37 weeks), mode of delivery, gestational age during delivery, birth weight (in grams), sex of the newborn, type of anesthesia during delivery) and clinical factors(history of preeclampsia/eclampsia, history of gestational diabetes, history of placenta previa, history of uterine leiomyomas, history of uterine structural abnormality and mode of conception) and dependent variable was placenta previa (Yes/No)

#### Data Collection Tool and Procedures

The data for this study was secondary data and were obtained from chart of pregnant women with the diagnosis of placenta previa (cases) and those without PP (controls) who admitted and managed from July 8, 2020 to June 30, 2023. All necessary (socio demographic factors, obstetric factors, and clinical factors) data for this research were extracted from the patient’s intake form, medical history registration book, any electronic data sources; clinical records, including laboratory results of biomarkers; and any relevant investigation using a structured and pretested questionnaire developed based on the maternal antenatal care follow-up chart applied in the hospital and different reviewed literature conducted previously [3, 4, 7, 25] in the English language. HMIS card numbers were utilized to identify individual patient cards or their data in the electronic database. The data were collected by health workers working at maternal and child health care follow-up unit who were carefully selected based on their experience and educational level. Data collectors and supervisor were received a two days training on the overall data collection procedure to develop a common understanding. Data was collected by two BSc midwives and was supervised by one general practitioner.

#### Data Quality Control

To ensure the data quality, an appropriately designed and pretested data extraction checklist was used, and training and necessary explanations for the data collector about the objectives of the study and process of the data collection were given. Every day, the collected data were checked for completeness and consistency. Familiarization with the checklist and strict supervision were provided during the data collection. The overall data collection process was controlled by principal investigator throughout the data collection period. The completed structured questionnaires was coded and entered into Epi-data manager version 4.6 for data exploration and cleaning.

#### Data Management and Analysis

After coding the questionnaire the data were entered into Epi-data manager version 4.6 for cleaning and exported to SPSS version 25 for statistical analysis. First, descriptive statistics were computed and the result was presented using frequencies and percentages. Then, bivariate and multivariable logistic regression was conducted and the presence of statistically significant association between the explanatory and outcome variables. Association was declared at P-value <0.05 and adjusted odds ratio (AOR) with 95% confidence interval. Multi-collinearity test was conducted using tolerance with (greater than 0.1) and variance inflation factor (VIF) at less than 10 and no collinearity exists between predictor variables. The model goodness of the test was checked by Hosmer-Lemeshow goodness of the fit and the P-value of the model fitness of the test was 0.752.x^2^ =3.44.

#### Ethical approval

Ethical approval and a letter of cooperation were obtained from the Institutional Review Board of Wolaita Sodo University, College of Medicine and Health Sciences with ref no.CRC SD/132/02/14 and all hospital were informed about the study objectives through a written letter. The formal letter of permission was written to the respective hospitals and the health management information system focal persons who verbally consented to the use of the patient’s medical charts for data collection. Informed consent was waived by the Wolaita Sodo University. Confidentiality was maintained at all levels of the study. The data were stored on a secured password protection system. All procedures were conducted based on the regulations, guidelines and principles of the Helsinki Declaration.

#### Patients and public involvement

Patients and the public were not involved in the design of the study, the conduct of the study or the dissemination of the findings.

## Results

### Socio-demographic Characteristics

Out of 19,677 deliveries, four hundred thirty one pregnant women delivered in study settings with 144 cases and 287 controls were included in this study. The age of study unit was ranged from 18-40 years, with the mean age of 27.35±5.04 years for both cases and controls. Both cases and controls, nearly a half 77/144 (53.5%) and 147/287(51.2%) were found in the age group of 25_34 years for cases and controls respectively (**Table 1**).

**Table 1.**
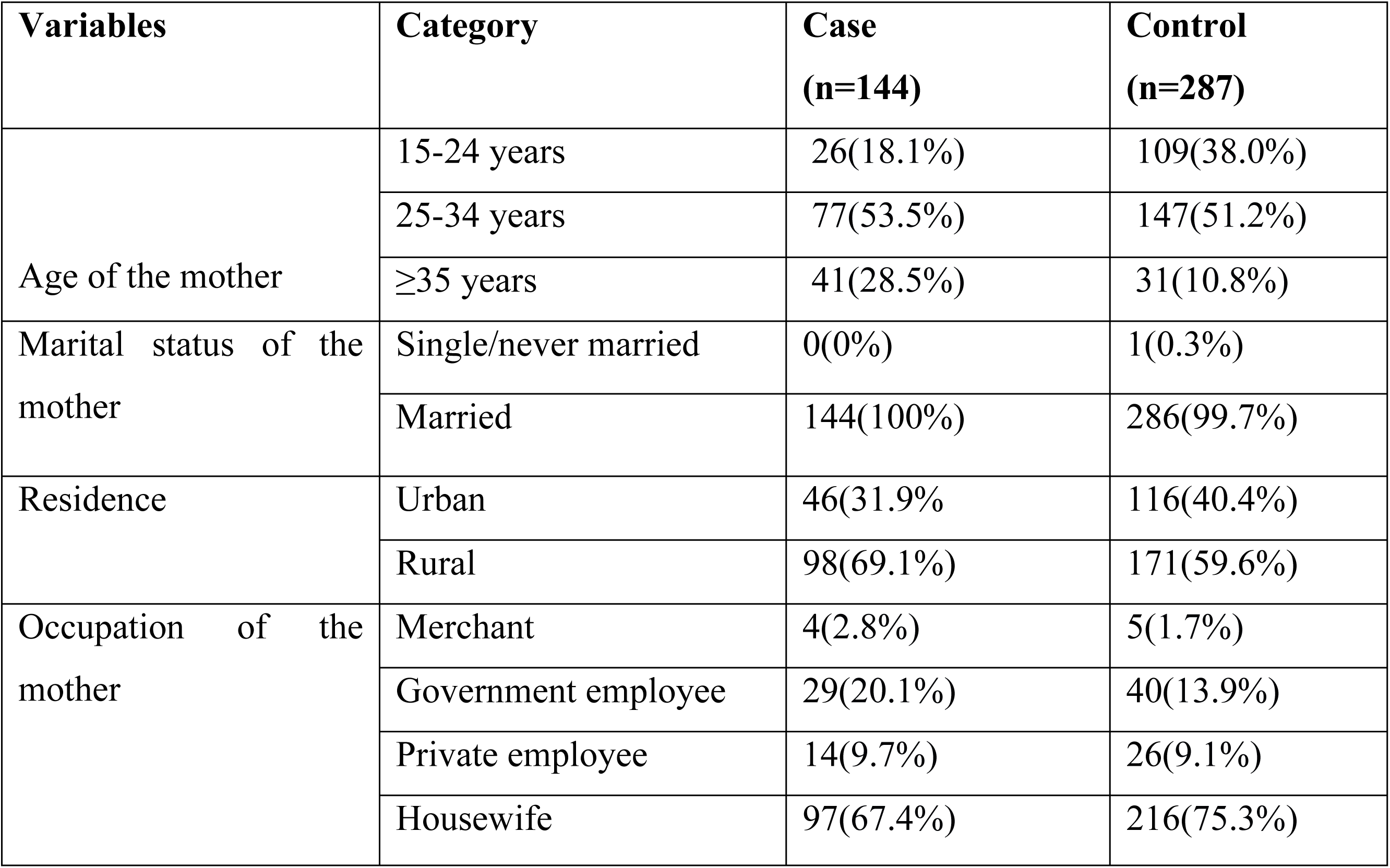
Socio-demographic Characteristics of Cases and Controls in public hospitals in the Wolaita zone southern regions of Ethiopia.

| **Variables** | **Category** | **Case**<br>**(n=144)** | **Control**<br>**(n=287)** |
| --- | --- | --- | --- |
| Age of the mother | 15-24 years | 26(18.1%) | 109(38.0%) |
|  | 25-34 years | 77(53.5%) | 147(51.2%) |
|  | ≥35 years | 41(28.5%) | 31(10.8%) |
| Marital status of the mother | Single/never married | 0(0%) | 1(0.3%) |
|  | Married | 144(100%) | 286(99.7%) |
| Residence | Urban | 46(31.9%) | 116(40.4%) |
|  | Rural | 98(69.1%) | 171(59.6%) |
| Occupation of the mother | Merchant | 4(2.8%) | 5(1.7%) |
|  | Government employee | 29(20.1%) | 40(13.9%) |
|  | Private employee | 14(9.7%) | 26(9.1%) |
|  | Housewife | 97(67.4%) | 216(75.3%) |

### Obstetric History of Current Pregnancy

Nearly half of cases found in pregnancy with gravida of 3_4 was 65/144 (45.1%), followed by gravida of ≥5, 42(29.2%) and gravida 1 to 2 37(25.7%). Out of 287 controls, nearly half 159(55.4%) was found in the pregnancy of gravida 1 and 2, which followed by gravida 3 and 4,102 (35.5%), and gravida ≥5, 26(9.1%). Out of 144 cases, nearly half was found in the para3 and 4 which was 62(43.1%)whereas among 287 controls, nearly 2/3^rd^ was found in the para 1 and 2,171(60.0%). Among 144 cases, nearly 2/3^rd^ 93 (64.6%) of the pregnant women delivered with short inter-provenance interval of less than 24 months and 36(3.4%) of the pregnant women were delivered inter-provenance interval of 24 months and above (**Table 2**).

**Table 2.**
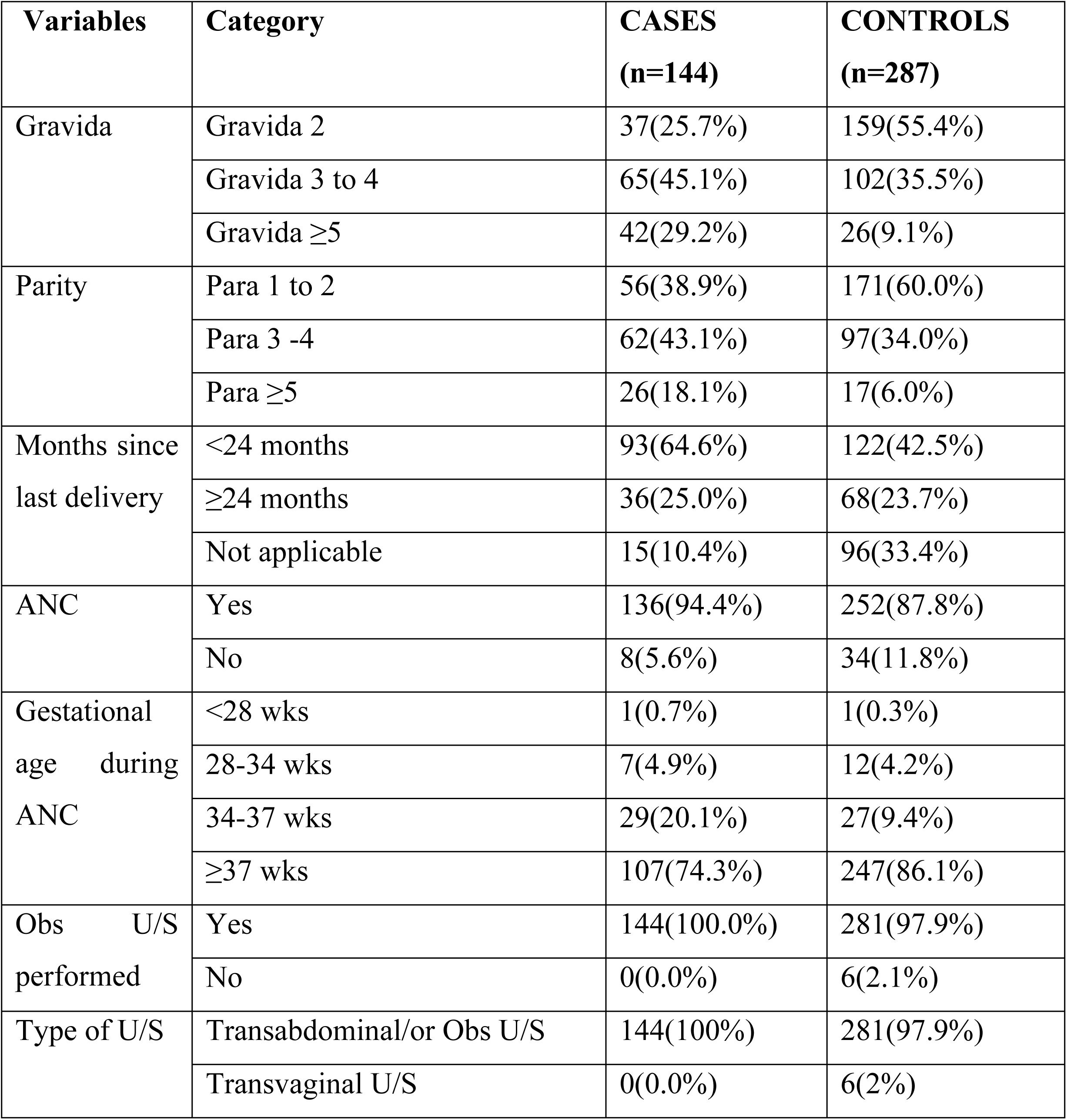
Obstetric Characteristics for Current Pregnancy of Cases and Controls in public hospitals in the Wolaita zone southern regions of Ethiopia.

| **Variables** | **Category** | **CASES**<br>**(n=144)** | **CONTROLS**<br>**(n=287)** |
| --- | --- | --- | --- |
| Gravida | Gravida 2 | 37(25.7%) | 159(55.4%) |
|  | Gravida 3 to 4 | 65(45.1%) | 102(35.5%) |
| | Gravida $\geq 5$ | 42(29.2%) | 26(9.1%) |
| Parity | Para 1 to 2 | 56(38.9%) | 171(60.0%) |
|  | Para 3 -4 | 62(43.1%) | 97(34.0%) |
| | Para $\geq 5$ | 26(18.1%) | 17(6.0%) |
| Months since last delivery | <24 months | 93(64.6%) | 122(42.5%) |
| | $\geq 24$ months | 36(25.0%) | 68(23.7%) |
|  | Not applicable | 15(10.4%) | 96(33.4%) |
| ANC | Yes | 136(94.4%) | 252(87.8%) |
|  | No | 8(5.6%) | 34(11.8%) |
| Gestational age during ANC | <28 wks | 1(0.7%) | 1(0.3%) |
|  | 28-34 wks | 7(4.9%) | 12(4.2%) |
|  | 34-37 wks | 29(20.1%) | 27(9.4%) |
| | $\geq 37$ wks | 107(74.3%) | 247(86.1%) |
| Obs U/S performed | Yes | 144(100.0%) | 281(97.9%) |
|  | No | 0(0.0%) | 6(2.1%) |
| Type of U/S | Transabdominal/or Obs U/S | 144(100%) | 281(97.9%) |
|  | Transvaginal U/S | 0(0.0%) | 6(2%) |

### Obstetric Characteristics

Out of 144 cases, nearly 1/3^rd^ had history of previous abortions, 64/144(44.4%), and nearly 2/3^rd^ had no history of previous abortions, 80/144(55.6%). Out of 287 controls, majority had no prior abortions history, 247/287(86.1%), and nearly 1/10^th^ had previous history of abortions, 40/287(13.9%). Out of 144 of the cases, 28(19.4%) had previous history of cesarean sections and in out of 287 of controls, 19(6.6%) had previous history of C/S. Out of 144 cases, majority of the current pregnancy was delivered by C/S, 113(78.5%), followed by SVD 31(21.5%) (**Table 3**).

**Table 3:**
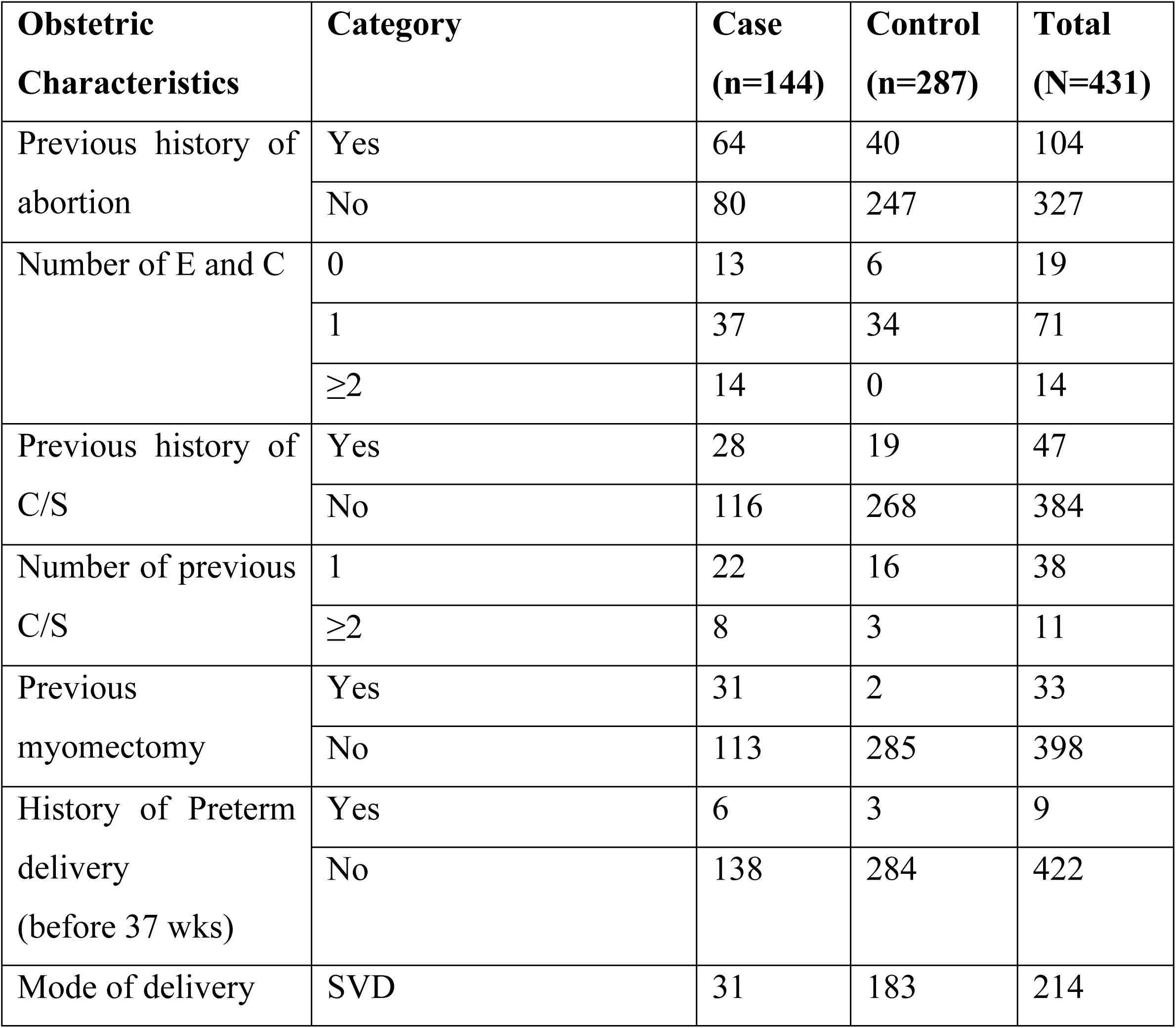
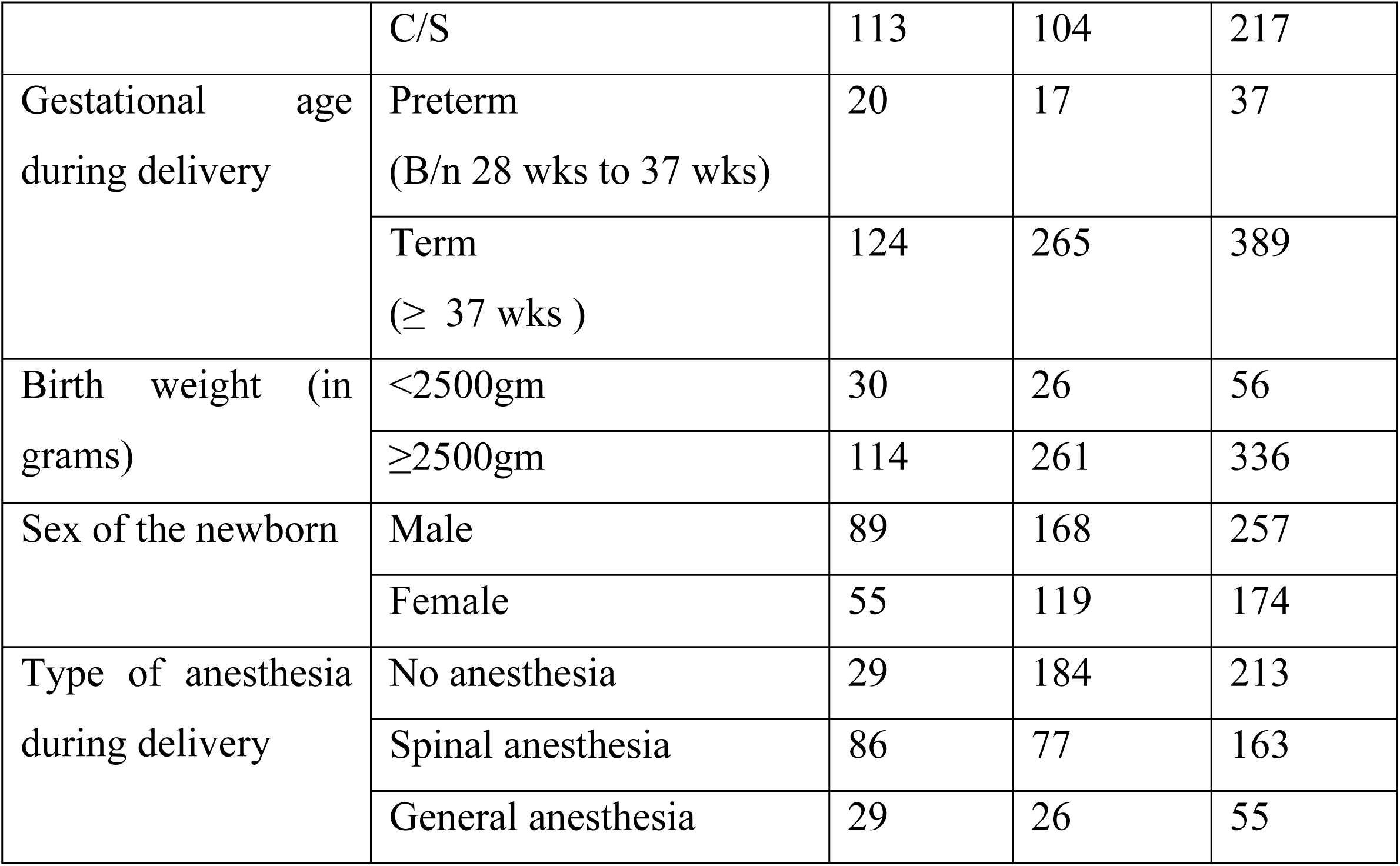
Obstetric Characteristics of cases and controls in public hospitals in the Wolaita zone southern regions of Ethiopia.

### Clinical Characteristics

Previous history of pre eclampsia or Eclampsia accounts 4(2.8%) out of 144 cases and 37(12.9%) out of 287 controls. Out of 144 cases, 22(15.3%) had previous history of placenta previa and from 287controls, history of placenta previa accounts 29(6.7%). History of uterine leiomyomas accounts 21(14.6%) from 144 cases and 16(5.6%) from 287 controls (**Table 4**).

**Table 4:** Clinical Characteristics of the Cases and Controls in public hospitals in the Wolaita zone southern regions of Ethiopia.

| Clinical Characteristics | Category | Cases<br>(n=144) | Controls<br>(n=287) |
| --- | --- | --- | --- |
| History of preeclampsia/eclampsia | Yes | 4(2.8%) | 37(12.9%) |
|  | No | 140(97.2%) | 250(87.1%) |
| History of gestational diabetes | Yes | 1(0.7%) | 1(0.3%) |
|  | No | 143(99.3%) | 286(99.7%) |
| History of placenta previa | Yes | 22(15.3%) | 29(6.7%) |
|  | No | 122(84.7%) | 280(97.6%) |
| History of uterine leiomyomas | Yes | 21(14.6%) | 16(5.6%) |
|  | No | 123(85.4%) | 271(94.4%) |
| History of uterine structural abnormality | Yes | 0(0.0%) | 4(1.4%) |
|  | No | 144(100%) | 283(98.6%) |
| Mode of Conception | Natural | 144(100%) | 287(100%) |
|  | ART | 0 | 0 |

### Bi-variate and Multi-variable Logistic Regression Analysis

Maternal age of ≥35 years, rural residence, short inter-pregnancy interval of <24 months, gravidity of ≥5, parity of ≥ 5, history of abortions, history of previous C/S, gestational age during delivery, mode of delivery by C/S, birth weight of below 2500gm, prior placenta previa and presence of fibroid s were statistically significant variables using bi-variate logistic regression and exported to multi-variable logistic regression.

Accordingly, maternal age of 35 years and above, having next pregnancy before 2 years (short inter-pregnancy period), having previous history of abortion, presence of previous cesarean sections, and pregnancy with the presence of uterine leiomyoma were identified determinants of placenta previa by using multi-variable logistic regression analysis.

The odds of developing PP was 4.45 among pregnant women with the age of 35 years and above compared to the maternal age less than 35 years [AOR=4.45 (95% C.I =1.2,, 4.99)]. The odds of developing PP was 1.89 among the pregnant women with the pregnancy of short inter-pregnancy interval of less than 24 months compared to the women of inter-pregnancy interval of 24 months and above [AOR=1.89 (95% C.I=(1.08, 3.53)]. The odds of developing PP was 5.49 among mothers with previous abortions when compared to the women with no prior abortions [AOR=5.49 (955 C.I =2.93, 10.3)]. The odds of developing PP was 3.73 among mothers with previous cesarean sections when compared to mothers without prior C/S [AOR=3.73 (95% C.I =1.68, 8.3)]. The odds of developing PP 6.33 was among women with women having uterine leiomyoma (fibroid s) when compared to women without uterine leiomyoma (fibroids) [AOR=6.33 (95% C.I=2.48, 16.17)]. The odds of developing PP was 4.72 among the pregnant women delivered low birth weight (<2500gm) when compared to the pregnant women delivered normal birth weight (≥2500gm) [AOR=4.72 (95% C.I=1.83, 12.17)]. Here, the relationship between low birth weight and placenta previa was about the neonatal effect of placenta previa rather than determinant factor (**Table 5**).

**Table 5:** Bi-variate and Multi-variable Logistic Regression for Determinants of Placenta Previa among Pregnant Women Delivered in public hospitals in the Wolaita zone southern regions of Ethiopia.

| Predictor variables | Category | Case (144) | Control (287) | COR(95%CI) | AOR (95%CI) |
| --- | --- | --- | --- | --- | --- |
| Maternal age | < 35 yrs | 103 | 256 | 1 | 1 |
| | ≥35 years | 41 | 31 | 3.29(1.96, 5.53) | 4.45*(1.2, 4.99) |
| Residence | Urban | 46 | 116 | 1 | 1 |
|  | Rural | 98 | 171 | 1.45(0.95, 2.20) | 1.33 (0.76, 2.35) |
| Gravidity | <5 | 102 | 261 | 1 | 1 |
| | ≥5 | 42 | 26 | 4.13(2.41, 7.09) | 1.58 (0.47, 5.29) |
| Parity | <5 | 118 | 270 | 1 | 1 |
| | ≥5 | 26 | 17 | 3.5(1.83,6.69) | 1.45(0.37, 5.66) |
| Months since last delivery | <24month | 93 | 122 | 2.47(1.63, 3.73) | 1.89*(1.08, 3.53) |
| | ≥24month | 51 | 165 | 1 | 1 |
| Abortion History | Yes | 64 | 40 | 4.94(3.09,7.89) | 5.49*** (2.93, 10.3) |
|  | No | 80 | 247 | 1 | 1 |
| History of C/S | Yes | 28 | 19 | 3.4(1.83, 6.34) | 3.73**(1.68, 8.3) |
|  | No | 116 | 268 | 1 | 1 |
| Mode of delivery | SVD | 31 | 183 | 1 | 1 |
|  | C/S | 113 | 104 | 6.4(4.03,10.21) | 6.55*** (3.75, 11.43) |
| Birth weight (in grams) | <2500gm | 30 | 26 | 2.64(1.49, 4.67) | 4.72**(1.83,12.17) |
|  | ≥2500gm | 114 | 261 | 1 | 1 |
| Prior placenta previa | Yes | 22 | 15 | 3.27(1.64, 6.52) | 2.38(0.96,5.9) |
|  | No | 122 | 272 | 1 | 1 |
| Uterine Leiomyoma | Yes | 21 | 16 | 2.89(1.46, 5.73) | 6.33*** (2.48, 16.17) |
|  | No | 123 | 271 | 1 | 1 |

Hosmer-Lemeshow goodness of-fit and the P-value of the model fitness of the test=0.75, $\chi^2$ (Chi-square)=3.44, COR: Crude Odds Ratio, AOR: Adjusted Odds Ratio, C.I: Confidence Interval, \*statistically significant at P-value <0.05, \*\*statistically significant at P-value <0.01, \*\*\*statistically significant at P-value <0.001

## Discussion

The study findings highlight significant determining factors to placenta previa, including increased maternal age (≥35 years), short inter-pregnancy interval of less than 24 months, history of abortions, prior cesarean sections and having leiomyoma.

In this study, the odds of developing placenta previa was 4.45 fold among the maternal age of 35 years and above compared to the maternal age of below 35 years. This study finding agree with other studies conducted previously, which states that the development of placenta previa was strongly associated with increased maternal age (especially >34 years) [5.9, 26-28]. The study conducted by Cleary shown that advancing maternal age increases the risk of placenta previa, those older than 35 years had a 1.1% risk for previa compared with that of 0.5% for women less than 35 [26]. According to a meta-analysis shown that maternal age above 34 years was strongly associated with the development of PP [27]. This study and other previous studies have shown that there is the association between the development of placenta previa and advanced maternal age. But there is no clearly approved mechanisms that show how the advanced maternal age impair the normal implantation of placenta. However, according to a population-based, case-control study explained the possible reason as atherosclerotic changes on uterine arteries increase with increasing maternal age, resulting in reduced blood supply and infarction of the placenta, thereby increasing the placental size, and hypertrophied and enlarged placenta increases the risk of the placental implantation on lower uterine segment [28, 29].

In this study, short inter-pregnancy interval of less than 24 months was significantly associated with the development of PP. The odds of developing PP was 1.89 among short inter-pregnancy interval of less than 24 months compared to the women being pregnant of inter-pregnancy interval of 24 months and above. This study finding supported with the study of conducted on association of short inter-pregnancy interval with pregnancy outcomes according to maternal age showed that women who have short inter-pregnancy interval of less than 18 months have been associated with an increased risk of adverse perinatal outcomes, including placenta previa [30]. However, this study finding is not in-line with the study conducted among Norwegian women, revealed that long inter-pregnancy interval of more than 4 years was associated with placenta previa. The exact mechanism is unknown, but most possible explanation could be due to uterine scarring or poor vasculature of the uterus which is associated with advanced maternal age [31]. The presence of strong association between short inter-pregnancy intervals of 24 months is not clearly identified; therefore, it needs further study.

The odds of developing placenta previa were 5.49 among pregnant women with previous abortions when compared to the women without previous abortions. Similarly, a meta-analysis study conducted revealed that previous history of abortions were significantly increase the risk of placenta previa development [32]. Another meta-analysis done reported that presence of previous abortions history would increase the risk of placenta previa occurrence and the risk of developing placenta previa was significantly associated with prior abortions [33]. Also, a matched case control study conducted in 2015 shown that prior abortion was significantly associated with placenta previa and the odds of having placenta previa were 2 times more likely in women having previous history of abortion [34]. There was no clear mechanism that explains how previous abortions predetermine the occurrence of placenta previa. However, the most possible explanations could be the damage and scarring of myometrium and endometrium of the uterus during repeated abortions may distort normal fundal implantation of the placenta, which in turn influences implantation of placenta at low uterine segment in the next pregnancies [31, 35].

The odd of developing placenta previa was 3.73 among pregnant women with previous cesarean section when compared to the pregnant women without prior cesarean section. This study finding is in line with the unmatched Case-Control Study conducted demonstrated that pregnant women who had previous cesarean section have 3 times increased risk of developing placenta previa [4]. Similarly, a matched case control study reported that previous history of cesarean section had statistically significant association with placenta previa [34]. Previous several studies have reported an association between placenta previa and previous history of cesarean sections. Those studies were also demonstrated that previous history of cesarean section increased the risk of developing placenta previa [9, 30, 32, 36]. This is due to the surgical damage to the uterus of endometrial and myometrial lining and causing the formation of the endometrial scarring. Once a previous cesarean section is done on the endomyometrium of the uterus, angiogenesis is developed in the previous scar site that may result in partial hypoxia. Hypoxia causes incomplete decidualization and abnormal trophoblast invasion that may result the attraction and adherence of placental tissue to the lower uterine segment in subsequent pregnancies [33].

According to this study finding, there was high statistically significant association between low birth weight (birth weight of below 2500gm) and placenta previa. Here, the relationship between low birth weight and placenta previa was about the neonatal effect of placenta previa rather than determinant factor [21]. According to a population based study conducted women with PP were nineteen times more likely to deliver low birth weight (less than 2500gm) babies. The clear mechanism is unknown, but the most possible reason could be due to severity of bleeding that leads to premature delivery. Chronic hypoxia due to PP also results intra-uterine growth restriction and this could leads to the delivery of low birth weight neonate [37].

In this study finding, there was strong relationship between uterine leiomyoma (fibroid) and placenta previa. The odds of developing PP were 6.33 among the women of having uterine leiomyoma compared to those without leiomyoma. This study supported with other studies conducted which revealed that the presence of fibroid is associated with a 2-fold increased risk of PP even after adjusting for prior operations such as cesarean sections or myomectomy [38, 39]. Another similar report, done showed that here was significant association between uterine leiomyoma and placenta previa and the risk of developing PP was 2.21 times among women of having uterine leiomyoma compared to those without fibroid. This similar study mentioned that uterine leiomyoma of size 5cm or more increases the risk of placenta previa by 3.53 -fold [40]. There is no clearly known mechanism that explaining the association between fibroid and abnormal implantation of the placenta in the lower uterine segment. The most possible reason could be due to its highest occurrence related with increased maternal age, which result in reduced blood supply and infarction of the placenta, thereby increasing the placental size, and hypertrophied and enlarged placenta increases the risk of the placental implantation on lower uterine segment [28, 29].

### Limitations of the Study

Due to its facility based historical nature of the study, some incomplete and missed variables were excluded from the study during data collection. Again, the variables with small sample size that make other variables unstable were excluded during data analysis based on minimum sample size requirement. Therefore, its results may not be generalizable to the whole pregnant women of the Ethiopia.

## Conclusions

In this study, the determinants of placenta previa among pregnant women by unmatched case-control study were increased maternal age of 35 years and above, pregnancy with short inter-pregnancy interval of less than 24 months, mode of delivery through C/S, previous history of abortions, previous history of cesarean section, and presence of uterine leiomyoma (fibroids) were strongly associated with the development of placenta previa by multi-variable logistic regression analysis after adjusting other confounder factors like high parity, having more than 5 gravida, rural residence, and prior placenta previa. In this study finding the presence of association between low birth weight and placenta previa was not explained about risk factor, rather than the low birth weight was the effect of placenta previa. There should be a clear strategy and policy to identify determinants of placenta previa.

## Acronyms

AOR: Adjusted Odds Ratio
APH: Antepartum Hemorrhage
C/S: Caesarean Section
E and C: Evacuation and curation
PPH: PostPartum Hemorrhage
PP: Placenta previa
SVD: Spontaneous Vaginal Delivery

## Consent for publication

Not applicable

## Data availability statement

The data will be available upon request from the corresponding author.

## Author’s Contributions

TU: Conceptualization, Data curation, Formal analysis, Funding acquisition, Investigation, Methodology, Project administration, Resources, Software, Supervision, Validation, Visualization, Writing—original draft, Writing—review and editing;EW:Conceptualization, Data curation, Formal analysis, Funding acquisition, Investigation, Methodology, Project administration, Resources, Software, Supervision, Validation, Visualization, Writing—original draft, Writing—review and editing: TY: Conceptualization, Writing—original draft, Writing—review and editing;: Software, AA - Supervision, Validation, Visualization, Writing—original draft, Writing—review and editing: GK: Conceptualization, Validation, Visualization, Writing—original draft, Writing—review and editing:.JG - Conceptualization, Writing—original draft, Writing—review and editing;: Software.

## Acknowledgments

We would like to acknowledge Wolaita Sodo University, Graduate Studies Directorate, College of Medicine and Health Science, School of Public Health for their support during conducting this research.

